# Benchmarking And Datasets For Ambient Clinical Documentation: A Scoping Review Of Existing Frameworks And Metrics For AI-Assisted Medical Note Generation

**DOI:** 10.1101/2025.01.29.25320859

**Authors:** Sarah Gebauer

**Affiliations:** MachineLearningForMDs

**Keywords:** ambient scribe, artificial intelligence, clinical documentation, evaluation metrics, benchmarking, natural language processing

## Abstract

**Background:** The increasing adoption of ambient artificial intelligence (AI) scribes in healthcare has created an urgent need for robust evaluation frameworks to assess their performance and clinical utility. While these tools show promise in reducing documentation burden, there remains no standardized approach for measuring their effectiveness and safety.

**Objective:** To systematically review existing evaluation frameworks and metrics used to assess AI-assisted medical note generation from doctor-patient conversations, and provide recommendations for future evaluation approaches.

**Methods:** A scoping review following PRISMA guidelines was conducted across PubMed, IEEE Explore, Scopus, Web of Science, and Embase to identify studies evaluating ambient scribe technology between 2020-2025. Studies were included if they were peer-reviewed, focused on clinical ambient scribe evaluation from speaking to note production, and described an evaluation approach. Extracted data included evaluation metrics, benchmarking approaches, dataset characteristics, and model performance.

**Results:** Seven studies met inclusion criteria. Evaluation approaches varied widely, from traditional natural language processing metrics like ROUGE and BERTScore to domain-specific measures such as clinical accuracy and bias. Critical gaps identified include: 1) wide diversity of evaluation metrics making cross-study comparison challenging, 2) limited integration of clinical relevance in automated metrics, 3) lack of standardized approaches for crucial metrics like hallucinations and errors, and 4) minimal diversity in clinical specialties evaluated. Only two datasets were publicly available for benchmarking.

**Conclusions:** This review reveals significant heterogeneity in how ambient scribes are evaluated, highlighting the need for standardized evaluation frameworks. We propose recommendations for developing comprehensive evaluation approaches that combine automated metrics with clinical quality measures. Future work should focus on creating public benchmarks across diverse clinical settings and establishing consensus on critical safety and quality metrics.

## Introduction

### Overview

Artificial intelligence in healthcare has created new ways to solve old problems in healthcare such as the increasing documentation burden for physicians. For example, ambient scribes^1^, which can transcribe the discussion between a doctor and patient and organize that information into a formatted note, have become increasingly common.

There are increasing calls for measuring how well healthcare AI technology performs^2^. In other domains of generative AI and machine learning, datasets and benchmarks provide a way to evaluate the performance of AI systems. Datasets are sets of information such as question and answer (or prompt and response) pairs, which are sometimes incorporated into a benchmark. Benchmarks are composed of one or more datasets that are designed for evaluation^3^. There have been historical attempts to capture all healthcare-related benchmarks, but these are largely defunct^4^.

This study provides an overview of the types of evaluation approaches specifically related to ambient scribes that are used by researchers, discusses challenges in the current state of ambient scribe evaluation, and makes recommendations for future healthcare AI evaluation research.

### Current state of ambient scribe benchmarks

Common general benchmarks like the Massive Multitask Language Understanding^5^ (MMLU) provide a sense of the general knowledge of the AI systems, while domain-specific benchmarks, such as ClinicBench^6^ establish performance in one area of interest. Although there are flaws inherent in using automated benchmarks, such as difficulty interpreting their applicability to

real-world situations, they provide an objective comparison of the strengths and weaknesses of different AI systems. Most benchmarks and datasets in general AI evaluation are multiple choice questions, though auto-graded open-ended questions are increasingly being used^7^.

The Coalition for Healthcare AI (CHAI) and others have formed working groups to help health systems evaluate and implement the ambient scribe technology. These processes are consensus-based and necessarily take time to develop recommendations. These technical aspects are made more challenging by a lack of consensus about what constitutes “good”, “good enough” or “success”, and how performance on automated benchmarks translates to each of those categories. Opportunities exist for evaluation both at the stage of audio to transcription and transcription to note summarization, though these are rarely explicitly disentangled. In the absence of clear guidance, ambient scribe developers and researchers have used a variety of evaluation approaches, from established natural language processing (NLP) metrics to established human rating systems to the creation of entirely new metrics.

## Methods

Relevant papers were identified based on the PRISMA framework^8^ between January 1 and January 25, 2025 via PubMed, IEEE Explore, Scopus, Web of Science, and Embase. A description of the search terms is provided in Appendix A. Papers were included if they were in English, peer-reviewed, focused on clinical ambient scribe evaluation from the phase of speaking to note production, described an evaluation approach and were published between 2020 and 2025. Metanalyses, studies focused on education, preprints, un-refereed conference proceedings, and opinion pieces were excluded. Papers that evaluated the only effects of the AI scribe technology, such as the time spent by the physician or patient satisfaction, or described only automated speech recognition techniques or clinical note summarization techniques or evaluations, were excluded. The citations were compiled and duplicates were identified using Rayyan AI. Descriptions of selected benchmarks are included in Appendix A.

In total, fourteen possible papers were identified for possible inclusion. Three were excluded because they focused on data extraction from existing clinical notes, one was excluded because it focused on data summarization from existing clinical notes, one was excluded because it focused on annotation of clinical notes, and two were listed in the databases searched but as of this writing were not peer-reviewed.

Seven studies met inclusion criteria. The associated process is described in Figure 1. Descriptions of the included studies are included in Table 1.

**Table 1.** Description of included studies. The seven included studies were published between 2023-2024, reflecting the relatively new technology of ambient scribes. The studies were performed in a variety of settings and locations on two continents, with most studies focused on primary and specialty ambulatory care settings. The sample sizes varied considerably, from small pilots with 14 simulated encounters to large-scale implementations involving over 300,000 patient encounters. Most studies used simulated conversations rather than real patient encounters, likely due to privacy concerns.

| Study Name | Authors | Affiliation | Purpose of study | Year published | Description of data used | Benchmark verification |
| --- | --- | --- | --- | --- | --- | --- |
| Using ChatGPT-4 to Create Structured Medical Notes From Audio Recordings of Physician-Patient Encounters: Comparative Study<sup>9</sup> | Kernberg A, Gold JA, Mohan V | Oregon Health & Sciences University | Evaluate accuracy and quality of ChatGPT-4 generated SOAP notes from audio recordings | 2024 | 14 simulated patient-provider encounters representing various ambulatory specialties | Human expert validation of generated notes against physician-created rubrics; avg. 23.6 errors per case found |
| An Empirical Study of Clinical Note Generation from Doctor-Patient Encounters<sup>10</sup> | Ben Abacha A, Yim W, Fan Y, Lin T | Microsoft Health AI & Nuance Communications | Create benchmark dataset and evaluate summarization models for clinical notes | 2023 | MTS-DIALOG: 1,700 doctor-patient dialogues (16k turns) and clinical notes (6k sentences) | Human expert evaluation of generated summaries; automatic metrics (ROUGE, BERTScore, etc.) |
| Use of an ambient artificial intelligence tool to improve quality of clinical documentation<sup>11</sup> | Balloch J, Sridharan S, Oldham G, et al. | TORTUS AI & Great Ormond Street Hospital | Assess clinical utility of ambient AI tool for enhancing consultation documentation | 2024 | 47 simulated consultations across 8 specialties with professional actors | SAIL (Sheffield Assessment Instrument for Letters) scores by independent clinicians; NASA Task Load Index |

<sup>9</sup> Kernberg A, Gold JA, Mohan V. Using ChatGPT-4 to Create Structured Medical Notes From Audio Recordings of Physician-Patient Encounters: Comparative Study. *J Med Internet Res*. 2024;26:e54419. Published 2024 Apr 22. doi:10.2196/54419
<sup>10</sup> Asma Ben Abacha, Wen-wai Yim, Yadan Fan, and Thomas Lin. 2023. An Empirical Study of Clinical Note Generation from Doctor-Patient Encounters. In *Proceedings of the 17th Conference of the European Chapter of the Association for Computational Linguistics*, pages 2291–2302, Dubrovnik, Croatia. Association for Computational Linguistics.
<sup>11</sup> Jasmine Balloch, Shankar Sridharan, GERALYN Oldham, Jo Wray, Paul Gough, Robert Robinson, Neil J. Sebire, Saleh Khalil, Elham Asgari, Christopher Tan, Andrew Taylor, Dominic Pimenta, Use of an ambient artificial intelligence tool to improve quality of clinical documentation, *Future Healthcare Journal*, Volume 11, Issue 3, 2024, 100157, ISSN 2514-6645, https://doi.org/10.1016/j.fhj.2024.100157.

|  |  |  |  |  |  |  |
| --- | --- | --- | --- | --- | --- | --- |
| **Ambient Artificial Intelligence Scribes to Alleviate the Burden of Clinical Documentation**<sup>12</sup> | Tierney A, Gayre G, Hoberman B, et al. | Kaiser Permanente | Evaluate ambient AI scribe technology implementation | 2024 | 303,266 patient encounters across multiple specialties | Modified PDQI-9 scoring by experts; pilot results from 10,000 physician licenses |
| **The Ambient Clinical Intelligence Benchmark (ACI-Bench)**<sup>13</sup> | Wu Y, Lin S, et al. | Microsoft Nuance | Create comprehensive benchmark dataset for evaluating AI clinical note generation across different interaction modes | 2023 | 207 clinical encounters divided into three subsets: virtual assistant interactions, virtual scribe dictations, and natural doctor-patient conversations | Multiple automatic metrics (ROUGE, BERTScore, BLEURT, medcon) combined with human expert validation. ASR vs human transcription accuracy compared |
| **Evaluation of a Digital Scribe: Conversation Summarization for Emergency Department Consultation Calls**<sup>14</sup> | Sezgin E., Sirrianni J, Kranz K | Abigail Wexner Research Institute | Evaluate a proof-of-concept digital scribe system's performance in summarizing emergency department consultation calls | 2024 | 100 referral conversations among ED physicians | ROUGE, annotations for accuracy and recall |

<sup>12</sup> Aaron Tierney, Gregg Gayre, Brian Hoberman, Britt MAttern, Manuel Balleca, Patricia Kipnis, Vincent Liu, Kristine Lee. Ambient Artificial Intelligence Scribes to Alleviate the Burden of Clinical Documentation. *NEJM Catalyst*. doi: 10.1056/CAT.23.0404.
<sup>13</sup> Yim, Ww., Fu, Y., Ben Abacha, A. *et al.* Aci-bench: a Novel Ambient Clinical Intelligence Dataset for Benchmarking Automatic Visit Note Generation. *Sci Data* 10, 586 (2023). https://doi.org/10.1038/s41597-023-02487-3.
<sup>14</sup> Sezgin E, Sirrianni JW, Kranz K. Evaluation of a Digital Scribe: Conversation Summarization for Emergency Department Consultation Calls. *Appl Clin Inform*. 2024 May 15;15(3):600–11. doi: 10.1055/a-2327-4121. Epub ahead of print. PMID: 38749477; PMCID: PMC11268986.

|  |  |  |  |  |  |  |
| --- | --- | --- | --- | --- | --- | --- |
| Impact of a Digital Scribe System on Clinical Documentation Time and Quality: Usability Study<sup>15</sup> | van Buchem MM, Kant IMJ, King L, et al. | Leiden University Medical Center, University Medical Center Utrecht, Autoscriber B.V. | Investigate impact of Dutch digital scribe system on clinical documentation efficiency and quality | 2024 | 27 recordings of mock consultations with nonmedical individuals acting as patients based on 26 vignettes focused on chest pain presentations | Modified PDQI-9 scoring, ROUGE scores, BERTScore metrics, word count, lexical diversity analysis, PDQI-9<br>430 summaries evaluated by 22 medical students using 3 methods: manual, automated, and automated with editing |

**Figure 1.**
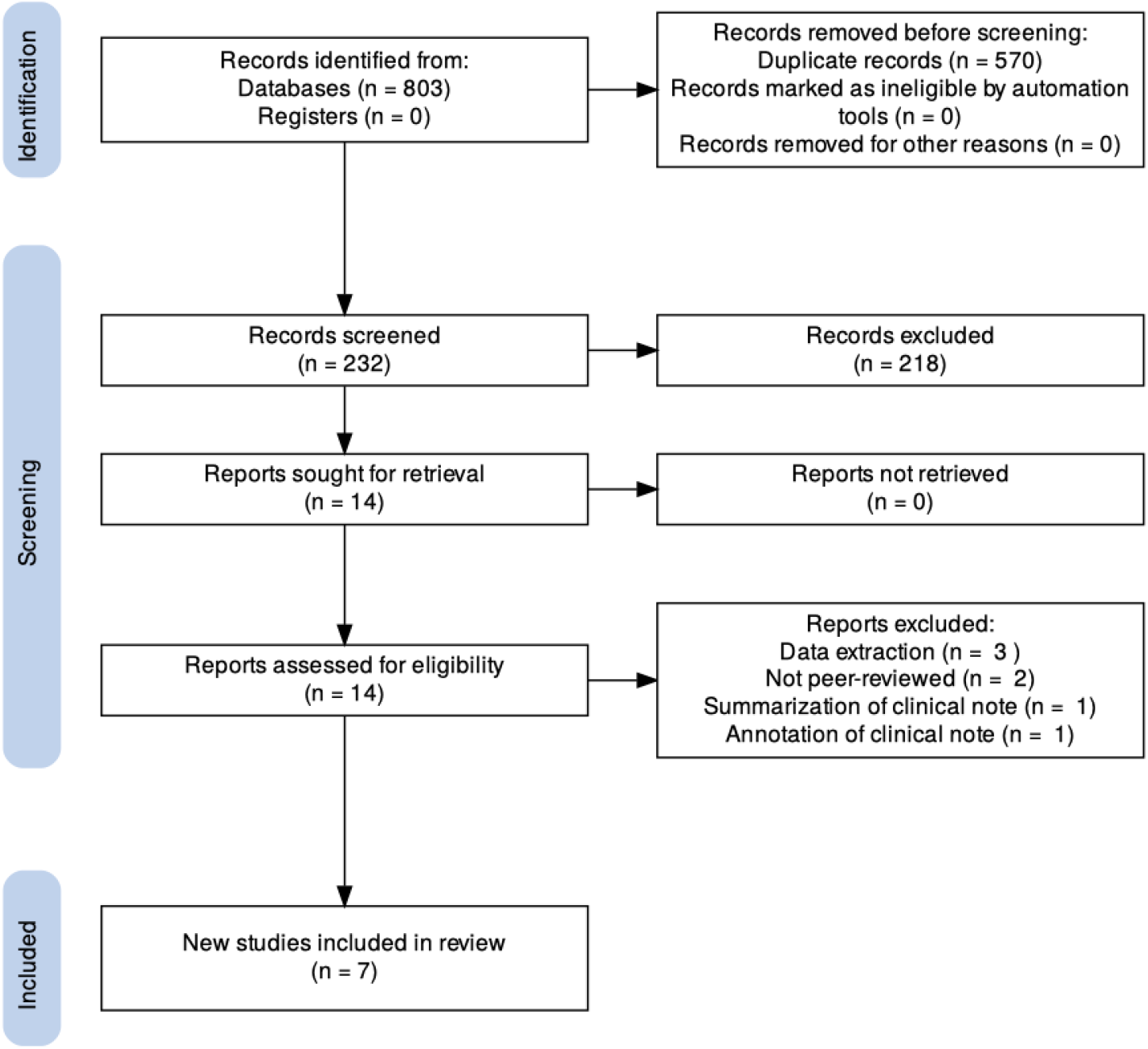
PRISMA diagram.

### Datasets used included studies

The datasets used in the included papers are described in the Appendix to provide the reader with information about the length of the dataset, amount of data, data source, specialty, and whether audio recordings are included in the dataset. They are listed here to provide readers with context for the metrics used, in that some used audio while others didn’t, and some were real conversations while others were simulated. These details provide clinicians with another layer of understanding regarding the metrics that are being reported. The datasets used varied substantially, from pre-recorded conversations, short snippets of dialogue, to real-time but minimally characterized conversations.As several of the papers associated with simulated datasets note, live conversations may be less focused as the visit often does not begin with a clear diagnosis in mind. Additionally, datasets without audio miss possible nuances related to accents and language barriers. Real conversations are challenging to release as public benchmarks due to concerns about patient privacy. MTS-DIALOG and ACI-Bench are publicly available.

**Table 2.** Datasets used in included studies.

| Study | Description | Real or simulated? | Specialty | Audio available? | Publicly available? |
| --- | --- | --- | --- | --- | --- |

<sup>15</sup> van Buchem MM, Kant IMJ, King L, Kazmaier J, Steyerberg EW, Bauer MP. Impact of a Digital Scribe System on Clinical Documentation Time and Quality: Usability Study. JMIR AI. 2024 Sep 23;3:e60020. doi: 10.2196/60020. PMID: 39312397; PMCID: PMC11459111.

|  |  |  |  |  |  |
| --- | --- | --- | --- | --- | --- |
| MTS-DIALOG | 1,700<br>conversations<br>(16k turns, 18k<br>sentences) | Simulated<br>conversations<br>created from<br>real clinical<br>notes | Multiple: General<br>Medicine, SOAP,<br>Neurology,<br>Orthopedic,<br>Dermatology,<br>Allergy/Immunology | No | Yes |
| Kaiser Dataset | 303,266 patient<br>encounters | Real | Multiple specialties<br>across<br>primary/specialty<br>care | No<br>(real-time<br>processing<br>only) | No |
| TORTUS Dataset | 47 simulated<br>consultations | Simulated with<br>professional<br>actors | 8 specialties:<br>Occupational<br>therapy, oncology,<br>cardiology,<br>speech/language<br>(cleft), respiratory,<br>urology,<br>dermatology,<br>speech/language<br>(feeding) | No | No |
| ChatGPT-4 Study<br>Dataset | 14 simulated<br>encounters | Simulated with<br>standardized<br>patients | Various ambulatory<br>specialties | Yes | No |
| ACI-Bench | 207 encounters<br>(55<br>turns/encounter<br>avg) | Simulated with<br>medical experts<br>and actors | Multiple specialties | Yes (for<br>some<br>subsets) | Yes |
| ED Consultation<br>Dataset | 100 referral<br>conversations | Real | Emergency<br>Department | Not<br>specified | No |
| Autoscriber<br>Dataset | 27 recordings of<br>mock<br>consultations | Simulated with<br>nonmedical<br>individuals | Primary care<br>(focused on chest<br>pain) | Yes | No |

### Models evaluated in included studies

The models evaluated directly impact both the datasets used and the results. Many studies ran tests on open-source models fine-tuned on medical data, with several testing versions of GPT as well. Models with short context lengths necessitate shorter conversational lengths, or performance is likely to suffer. The ACI-Bench study used a Longform Encoder Decoder (LED) to assist with BART’s context length limits. This list is helpful for researchers to identify commonly used models for evaluation. The Kaiser and Tortus studies used proprietary models and did not describe the underlying models; they are not included in the Table of Publicly Accessible Models in Appendix B.

## Results

### Evaluation approaches

Evaluation metrics can be grouped into a few general types:

1. N-gram based methods like ROUGE
2. Embedding-based metrics like BERT, COMET, and BLEURT
3. Fact-based evaluations like Fact-Core and Fact-Full
4. Clinical assessment with the PDQI-9 and SAIL methods
5. Computed metrics like hallucination rate and omission rate

Four of the seven studies evaluated text generation quality metrics, and five of the studies evaluated clinical accuracy metrics. Three of the studies used PDQI-9 quality metrics and another used SAIL metrics. MTS-Dialog employed the most diverse metrics with eight metrics. ROUGE metrics were the most common automated metrics, used in four studies.

Although quantitative NLP metrics and clinical metrics are complementary and could be performed together in many studies, few researchers reported both types of metrics. Some studies such as those done by Tortus AI and Kaiser reported additional metrics such as physician and patient satisfaction that fall outside the scope of this scoping review. Some studies reported metrics with similar approaches but named them differently, such as incorrect facts, additions, or hallucinations. Some studies used different metrics that were named similarly, such as accuracy.

**Table 3.**
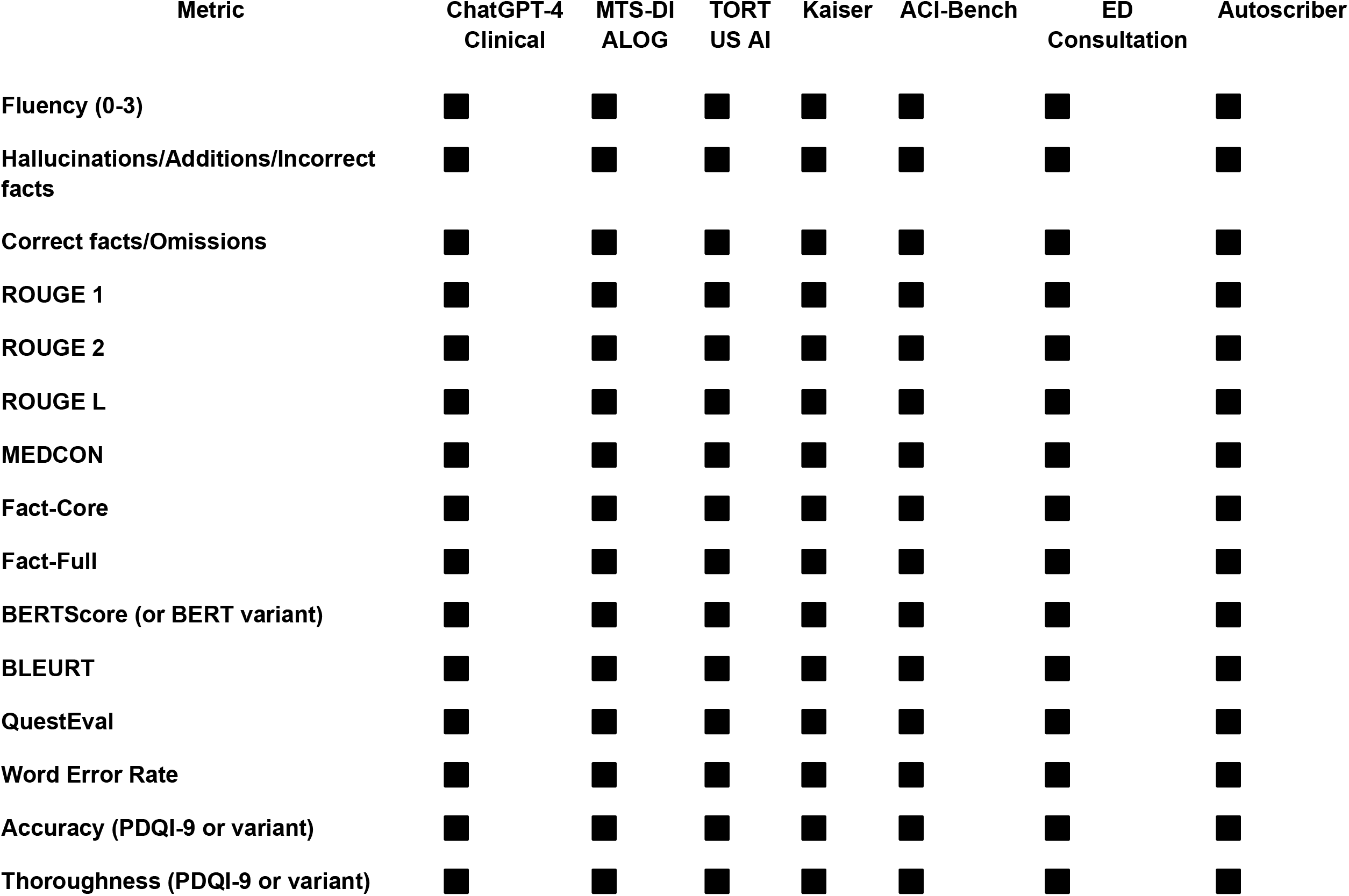
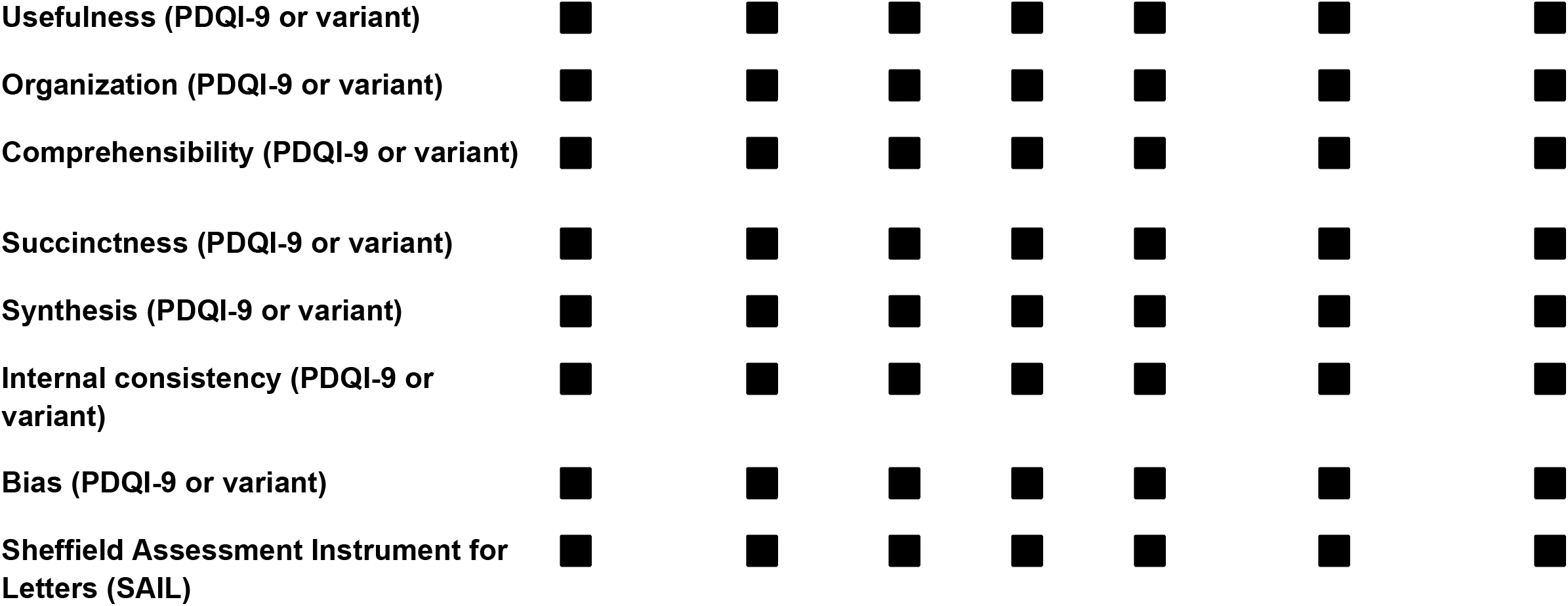
Metrics used in studies reviewed. Metric Used (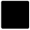= Present, 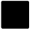= Absent)

### Limitations of this study

This review was meant to illustrate the current state of ambient scribe benchmarks and datasets related to evaluation of the software’s performance, but a more thorough systematic review that included additional metrics such as physician and patient satisfaction would likely provide additional information and details. This was performed by a single author; additional authors may have included additional studies, or excluded some of the included studies. The search terms were purposely limited to ambient scribes, but it is possible that more broad parameters may have identified additional studies and metrics. This study is also limited by publication bias, challenges in comparing different evaluation methods, and having a small number of studies.

## Discussion

### Diversity of evaluation approaches

Two major approaches to evaluation of AI scribes were identified: traditional NLP metrics (BLUERT, ROUGE) and clinical note-scoring frameworks (PDQI-9 with or without modification). Studies performed by clinicians tended to use subjective human measurements for potentially quantifiable aspects of evaluation. These aspects included accuracy, comprehensibility, internal consistency, and hallucinations.

Additionally, sometimes the same metric such, as accuracy, was measured either with traditional NLP approaches or with the subjective PDQI-9, but never with both approaches within a single study. These disparate methods make it challenging to compare performance across models and establish standards.

Even more metrics and approaches were identified in papers that have not undergone peer-review but cover similar material. For example, improving Clinical Note Generation from Complex Doctor-Patient Conversation^16^ describes the development of a dataset, CliniKnote, of 1,200 complex doctor-patient conversations with full clinical notes that are scored with automatic metrics (ROUGE, BERTScore, BLEURT) and human expert evaluation of keyword extraction. Similarly, DeepScore: A Comprehensive Approach to Measuring Quality in AI-Generated Clinical Documentation^17^ describes in a preprint how DeepScribe, an ambient scribe company, approaches metric generation including the creation of novel metrics such as critical error defect rate and the rate at which the clinician corrected the record. Another ambient scribe company, Tortus AI, developed novel metrics for hallucinations and clinical quality, described in their preprint^18^. In contrast, the primary outcome was solely of physician well-being in a preprint study^19^ of ambient scribe implementation at the University of Wisconsin. These preprint papers may go on to be published in a peer-reviewed format and be applicable for inclusion at that point; mentioning these papers and their included metrics is provided to emphasize the rapid pace of the field.

### The role of ambient scribe developers

Microsoft researchers wrote the ACI-Bench paper and the MTS-Dialogue paper. Other metrics that were not included in this review because the papers were not peer reviewed included the development of novel metrics such as a measurement of number of defects per note for a paper by DeepScribe, and an approach to evaluate hallucinations by Tortus AI. The contributions by ambient scribe developers suggest that these questions are active areas of investigation by industry, and that their expertise could be of assistance in discussions related to research and consensus-building.

### Description of grading approaches

Some studies focused on the fidelity of the audio to the transcription, while others focused more on the quality of the resulting note. Many studies did not report details about grading, particularly of human graders. Human graders may be influenced by incentives such as number of questions answered compared to number of questions answered correctly, or may have general expertise but limited expertise in a specific subject area. Inter-rater reliability was also generally not published. The automatically graded responses often did not provide details about whether these findings were confirmed with human spot-checking.

### Simulated conversations

While understandable from a privacy standpoint, many of the datasets are simulated both with regards to the cases that are presented for evaluation and the the conversations themselves. This makes contextualization and understanding application in a real-world environment more challenging.

### Minimal diversity in specialties

There were no simulations for the entire field of pediatrics, or for non-physician professionals. These users may have distinct needs for both transcription and note generation. For example, it is not known how well ambient scribes perform when toddlers describe their symptoms, or when a dietician needs to record brand names of specific goods. Additionally, many people may be speaking at once in many settings like the emergency room and current benchmarks do not evaluate these aspects.

### Experimental model setting reporting

Many papers failed to mention standard machine learning experimental settings like temperature and zero-shot vs few shot. These could contribute to different results even when using the same datasets and models. These make it even more challenging to compare models and studies against each other.

### Challenges with context window length

Conversations between physicians and patients are often long and include a large amount of information that may or may not be pertinent. Constraints related to short context lengths for the automated graders forced several workarounds and may have influenced performance.

### Recommendations for ambient scribe metrics

1. **Develop a standardized suite of ambient scribe metrics and datasets**
  a. The field should develop a set of metrics that uses quantifiable metrics when possible and incorporates clinical efficacy.
  b. Evaluations should distinguish between scoring at the audio to transcription level and the transcription to note organization levels.
  c. More publicly available evaluations of ambient scribes would make it easier to distinguish amongst top-performing models.
2. **Automate metrics when possible**
  a. Evaluation metrics need to be automatable whenever possible and repeatable, with minimal human oversight in order to simplify ongoing monitoring.
  b. For more subjective metrics, creating and validating public autograders that perform similarly to human experts would be a major contribution to the field and allow more rapid evaluation of metrics that are relevant and understandable to clinicians.
3. **Contextualize evaluation metrics**
  a. Baseline performance for current tools should be included when possible in order to contextualize performance.
  b. Metrics should be put in context of the current state when possible, as in using Automatic Speech Recognition and human transcription in several of the studies, or providing human baselines.
4. **Perform quality assurance**
  a. Evaluation metrics should be assessed for clinical correlation on an ongoing basis
  b. Evaluation metrics should be regularly checked to ensure grader models continue to align with desired outputs.
  c. Details about grading should be routinely reported, including details about the graders whether human or automated, the interrater reliability, and the incentives provided for training.
5. **Create evaluation approaches that anticipate likely advances in AI technology**
  a. As agentic models and models that interact with other models become more common, different approaches will be needed.
  b. If evaluations are only developed for the current technology, evaluation researchers will always be behind in creating public and private evaluation methods.
6. **Follow best practices for machine learning publications**
  a. Experimental settings such as temperature and zero-shot vs few-shot should be reported routinely.
  b. NeurIPS guidelines^20^ for data availability and reproducibility should be followed when possible.

## Conclusion

The field of AI system evaluations of healthcare AI, including ambient scribe systems, is still in its infancy, and there is minimal agreement amongst developers and researchers about which metrics should be measured. These issues will only become more challenging as the technology improves and agentic AI systems become common, and as hospitals and health systems implement multiple AI systems that interact with each other.

No one test is likely to answer all evaluation questions. A suite of evaluations that includes automated benchmarks, human-computer interaction studies, and interaction studies of multiple AI systems are more likely to produce meaningful evaluations. Many studies are time- and resource-intensive to develop, from creating high-quality samples and answers to using human experts to assess performance or serve as the gold standard. The faster, automated benchmarks can identify high-value questions and screen for emerging capabilities, while the slower and more expensive real-world simulations can provide more in-depth information.

The field needs a robust science of healthcare AI evaluations to provide the meaningful evidence-based metrics that clinicians can rely on. Future studies should pursue a more structured literature review to fully assess the field and as consensus develops from organizations like CHAI.

## Data Availability

All data referenced in the present work are publicly available at the referenced locations in the manuscript.

## Appendix

### Appendix A. Queries performed

#### Pubmed query: 37

((“ambient artificial intelligence” OR “ambient AI” OR “ambient scribe” OR “note generation” OR “automated documentation” OR “audio”) AND (“evaluation” OR “metrics” OR “assessment” OR “benchmark” OR “validation” OR “performance” OR “quality metrics”) AND (“note generation” OR “clinical documentation” OR “medical notes” OR “clinical notes”)) OR ((“large language model” OR “LLM” OR “transformer” OR “ChatGPT”) AND (“evaluation” OR “metrics”) AND (“clinical documentation” OR “medical transcription”))

#### IEE query: 2

((“ambient AI” OR “ambient artificial intelligence” OR “ambient scribe” OR “automated scribe” OR “autonomous documentation” OR “note generation” OR “large language model” OR “LLM” OR “transformer”) AND (evaluation OR metrics OR benchmark OR assessment OR validation OR performance OR “quality metrics”) AND (“clinical documentation” OR “medical notes” OR “clinical notes” OR “medical transcription” OR “healthcare documentation”))

#### Scopus query: 10

TITLE-ABS-KEY((“ambient AI” OR “ambient artificial intelligence” OR “ambient scribe” OR “automated scribe” OR “autonomous documentation” OR “note generation” OR “large language model” OR “LLM” OR “transformer”) AND (evaluation OR metrics OR benchmark OR assessment OR validation OR performance OR “quality metrics”) AND (“clinical documentation” OR “medical notes” OR “clinical notes” OR “medical transcription” OR “healthcare documentation”))

#### Web of Science query: 123

TS=((“ambient AI” OR “ambient artificial intelligence” OR “ambient scribe” OR “automated scribe” OR “autonomous documentation” OR “note generation” OR “large language model” OR “LLM” OR “transformer”) AND (evaluation OR metrics OR benchmark OR assessment OR validation OR performance OR “quality metrics”) AND (“clinical documentation” OR “medical notes” OR “clinical notes” OR “medical transcription” OR “healthcare documentation”))

#### Embase query: 109

(‘ambient artificial intelligence’ OR ‘ambient AI’ OR ‘ambient scribe’ OR ‘automated scribe’ OR ‘autonomous documentation’ OR ‘note generation’ OR ‘large language model’ OR ‘transformer’/exp) AND (‘evaluation’/exp OR ‘metrics’ OR ‘benchmark’ OR ‘assessment’ OR ‘validation’ OR ‘performance’ OR ‘quality metrics’) AND (‘clinical documentation’/exp OR ‘medical notes’ OR ‘clinical notes’ OR ‘medical transcription’ OR ‘healthcare documentation’)

### Appendix B. Descriptions of Selected Metrics

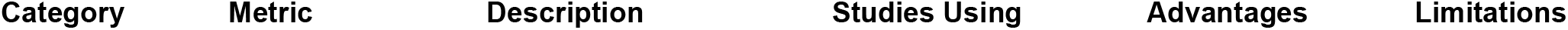
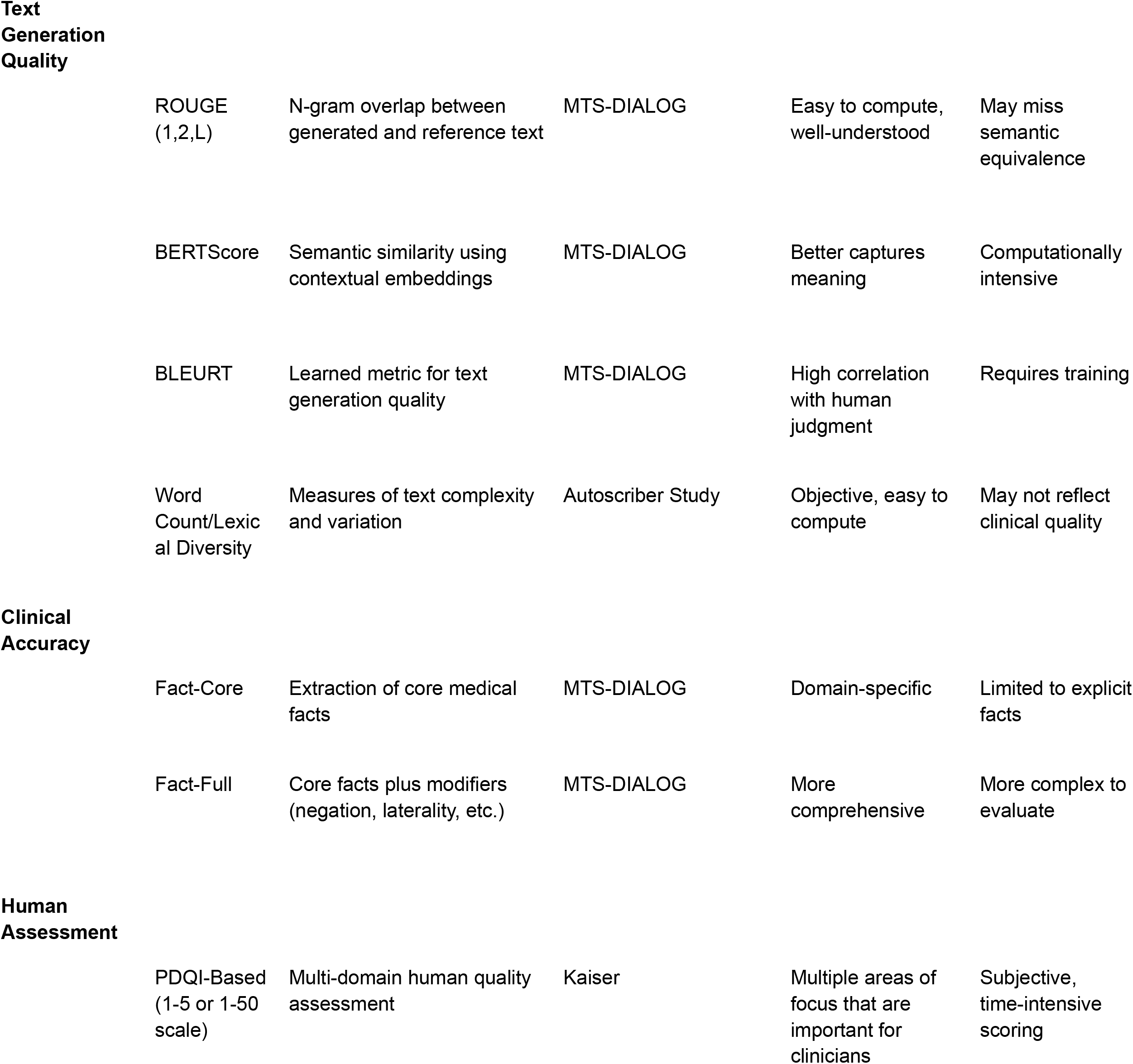
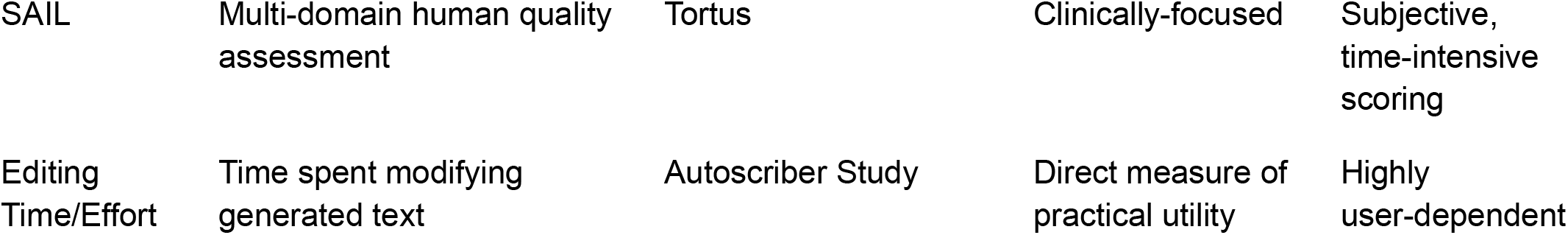

| Category | Metric | Description | Studies Using | Advantages | Limitations |
| --- | --- | --- | --- | --- | --- |

**Text Generation Quality**
|  |  |  |  |  |
| --- | --- | --- | --- | --- |
| ROUGE<br>(1,2,L) | N-gram overlap between<br>generated and reference text | MTS-DIALOG | Easy to compute,<br>well-understood | May miss<br>semantic<br>equivalence |
| BERTScore | Semantic similarity using<br>contextual embeddings | MTS-DIALOG | Better captures<br>meaning | Computationally<br>intensive |
| BLEURT | Learned metric for text<br>generation quality | MTS-DIALOG | High correlation<br>with human<br>judgment | Requires training |
| Word<br>Count/Lexical<br>Diversity | Measures of text complexity<br>and variation | Autoscriber Study | Objective, easy to<br>compute | May not reflect<br>clinical quality |

**Clinical Accuracy**
|  |  |  |  |  |
| --- | --- | --- | --- | --- |
| Fact-Core | Extraction of core medical<br>facts | MTS-DIALOG | Domain-specific | Limited to explicit<br>facts |
| Fact-Full | Core facts plus modifiers<br>(negation, laterality, etc.) | MTS-DIALOG | More<br>comprehensive | More complex to<br>evaluate |

**Human Assessment**
|  |  |  |  |  |
| --- | --- | --- | --- | --- |
| PDQI-Based<br>(1-5 or 1-50<br>scale) | Multi-domain human quality<br>assessment | Kaiser | Multiple areas of<br>focus that are<br>important for<br>clinicians | Subjective,<br>time-intensive<br>scoring |
| SAIL | Multi-domain human quality assessment | Tortus | Clinically-focused | Subjective, time-intensive scoring |
| Editing Time/Effort | Time spent modifying generated text | Autoscriber Study | Direct measure of practical utility | Highly user-dependent |

### Appendix C. Publicly accessible models evaluated in included studies

| Model Name | Size | Fine-tuned on what data? | Public? | Context Length | Studies that used this model |
| --- | --- | --- | --- | --- | --- |
| GPT-4 and other GPT versions | Unspecified | General | No | 32K | ChatGPT-4 Medical Notes Study, ACI-Bench |
| BART variants including BART-large | 400M (BART-large) | General | Yes | 1024 | MTS-DIALOG Study, ACI-Bench, ED Consultation Study, Autoscriber Study |
| BioBart | 400M | PubMed abstracts | Yes | 1024 | MTS-DIALOG Study, ACI-Bench |
| GPT-3.5-turbo | Unspecified | General + Clinical domain fine-tuning | No | 16K | ACI-Bench, Autoscriber Study |
| BERT-based (various) | 110M-340M | Various medical/clinical corpora | Yes | 512 | MTS-DIALOG Study (evaluation) |
| LED | Unspecified | General | Yes | 16K | ACI-Bench |
| LED+FT PubMed | Unspecified | PubMed | Yes | 16K | ACI-Bench |
| Text-davinci-002 and -003 | Unspecified | General + Clinical domain fine-tuning | No | 4K | ACI-Bench |
| T5-small and -large | Unspecified | General | Yes | Not specified | ED Consultation Study |
| Pegasus-PubMed | Unspecified | PubMed | Yes | Not specified | ED Consultation Study |

## Footnotes

1 Seth P, Carretas R, Rudzicz F. The Utility and Implications of Ambient Scribes in Primary Care. JMIR AI 2024;3:e57673. doi: 10.2196/57673

2 https://chai.org/

3 Kathrin Blagec, Jakob Kraiger, Wolfgang Frühwirt, Matthias Samwald, Benchmark datasets driving artificial intelligence development fail to capture the needs of medical professionals, *Journal of Biomedical Informatics*, Volume 137, 2023, 104274, ISSN 1532-0464, https://doi.org/10.1016/j.jbi.2022.104274

4 Kathrin Blagec, Jakob Kraiger, Wolfgang Frühwirt, Matthias Samwald, Benchmark datasets driving artificial intelligence development fail to capture the needs of medical professionals, *Journal of Biomedical Informatics*, Volume 137, 2023, 104274, ISSN 1532-0464, https://doi.org/10.1016/j.jbi.2022.104274

5 [2009.03300] Measuring Massive Multitask Language Understanding. Accessed 20th Jan 2025.

6 https://arxiv.org/pdf/2405.00716. Accessed 20th Jan 2025.

7 https://www.sciencedirect.com/science/article/pii/S0957417423011429. Accessed 28th Jan 2025.

8 Moher D, Liberati A, Tetzlaff J, Altman DG; PRISMA Group. Preferred reporting items for systematic reviews and meta-analyses: the PRISMA statement. PLoS Med. 2009 Jul 21;6(7):e1000097. doi: 10.1371/journal.pmed.1000097. Epub 2009 Jul 21. PMID: 19621072; PMCID: PMC2707599.

9 Kernberg A, Gold JA, Mohan V. Using ChatGPT-4 to Create Structured Medical Notes From Audio Recordings of Physician-Patient Encounters: Comparative Study. *J Med Internet Res*. 2024;26:e54419. Published 2024 Apr 22. doi:10.2196/54419

10 Asma Ben Abacha, Wen-wai Yim, Yadan Fan, and Thomas Lin. 2023. <u>An Empirical Study of Clinical Note Generation from Doctor-Patient Encounters</u>. In *Proceedings of the 17th Conference of the European Chapter of the Association for Computational Linguistics*, pages 2291–2302, Dubrovnik, Croatia. Association for Computational Linguistics.

11 Jasmine Balloch, Shankar Sridharan, Geralyn Oldham, Jo Wray, Paul Gough, Robert Robinson, Neil J. Sebire, Saleh Khalil, Elham Asgari, Christopher Tan, Andrew Taylor, Dominic Pimenta, Use of an ambient artificial intelligence tool to improve quality of clinical documentation, *Future Healthcare Journal*, Volume 11, Issue 3, 2024, 100157, ISSN 2514-6645, https://doi.org/10.1016/j.fhj.2024.100157.

12 Aaron Tierney, Gregg Gayre, Brian Hoberman, Britt MAttern, Manuel Ballesca, Patricia Kipnis, Vincent Liu, Kristine Lee. Ambient Artificial Intelligence Scribes to Alleviate the Burden of Clinical Documentation. *NEJM Catalyst*. doi: 10.1056/CAT.23.0404.

13 Yim, Ww., Fu, Y., Ben Abacha, A. *et al*. Aci-bench: a Novel Ambient Clinical Intelligence Dataset for Benchmarking Automatic Visit Note Generation. *Sci Data* 10, 586 (2023). https://doi.org/10.1038/s41597-023-02487-3.

14 Sezgin E, Sirrianni JW, Kranz K. Evaluation of a Digital Scribe: Conversation Summarization for Emergency Department Consultation Calls. Appl Clin Inform. 2024 May 15;15(3):600–11. doi: 10.1055/a-2327-4121. Epub ahead of print. PMID: 38749477; PMCID: PMC11268986.

15 van Buchem MM, Kant IMJ, King L, Kazmaier J, Steyerberg EW, Bauer MP. Impact of a Digital Scribe System on Clinical Documentation Time and Quality: Usability Study. JMIR AI. 2024 Sep 23;3:e60020. doi: 10.2196/60020. PMID: 39312397; PMCID: PMC11459111.

16 https://arxiv.org/html/2408.14568v1. Accessed 20th Jan 2025.

17 https://arxiv.org/abs/2409.16307. Accessed 20th Jan 2025.

18 https://www.medrxiv.org/content/10.1101/2024.09.12.24313556v1. Accessed 27th Jan 2025.

19 Afshar M, et al. A Novel Playbook for Pragmatic Trial Operations to Monitor and Evaluate Ambient Artificial Intelligence in Clinical Practice. medRxiv 2024.12.27.24319685; doi: https://doi.org/10.1101/2024.12.27.24319685. Accessed 26th Jan 2025.

20 https://arxiv.org/pdf/2003.12206. Accessed 20th Jan 2025.

## Notes

### Competing Interest Statement

Dr. Gebauer serves as a consultant to several healthcare AI companies including TORTUS AI in the past. None of these companies reviewed this manuscript, provided feedback, or were aware of it prior to publication.

### Funding Statement

This study did not receive any funding.

